# From Lectures to Learning Outcomes: Meaningful Integration of AI-Generated Content in Pre-Clerkship Medical Training

**DOI:** 10.1101/2025.05.13.25327518

**Authors:** Jay Khurana, Hossam A Zaki, Ellie Pavlick, Jillian Turbitt, Heather McGee, Sahil Gupta, Sriya Sai Pushpa Datla, Salma Eldeeb, Thais Salazar Mather, Sarita Warrier, Joyce Ou

**Author notes:** Corresponding Author Jay Khurana.

## Abstract

**Background:** Large Language Models (LLMs) can efficiently synthesize educational content, yet few studies have evaluated standardized, LLM-powered curricular interventions and their effects on medical student learning.

**Methods:** In this single-institution study, lecture-specific summaries and Anki flashcards for the pre-clerkship curriculum were generated using LLMs optimized through a structured, content-agnostic prompt engineering process. Outputs were evaluated using standardized human grading for (1) hallucinations, (2) coverage of faculty-defined learning objectives, and (3) flashcard information density. The highest-performing configurations produced materials deployed across two 3-week blocks (genetics and pharmacology). Impact was assessed using block exam performance and a post-intervention survey in a resource-rich environment with existing faculty-created, commercial, and student-curated materials. Because AI use was self-selected, we compared baseline performance on the three preceding block exams and tested for a catch-up effect using difference-in-differences and analysis of covariance (ANCOVA).

**Results:** Optimized prompts yielded zero hallucinations in summaries and 1 per 21 flashcards (4.8%), with 100% mean coverage of faculty-defined learning objectives. Exam performance did not differ significantly between summary users and non-users in either the genetics (p = 0.76) or pharmacology (p = 0.35) block, and between-group differences fell within a prespecified equivalence margin in adequately powered comparisons. Flashcard use was likewise not associated with significant score differences (genetics p = 0.86; pharmacology p = 0.05), while AI-flashcard users had lower baseline performance than non-users (Cohen’s d = -0.36 to -0.59). Most users reported time savings with AI flashcards (74%) and summaries (61%) and rated them more time-saving than the platform’s default GPT summary (91%); greater use correlated with higher perceived utility for faculty notes and student flashcards (r² = 0.55, 0.79).

**Conclusions:** Pre-clerkship students who used AI-generated content showed no significant difference in exam outcomes, with differences within a prespecified equivalence margin in adequately powered comparisons, while reporting time savings and usefulness; baseline data suggest these users began with lower prior performance and caught up, though small subgroups (e.g., pharmacology flashcards, n=14), regression-to-the-mean effect, and near-universal genetics-summary exposure (92 of 98) limit inference. AI-generated content may offer an efficiency for programs producing materials and an opt-in study option within a well-resourced curriculum. Future work will examine less structured settings, such as clinical and surgical education.

## Introduction

Medical education has traditionally relied on lecture-based teaching and self-directed study to prepare future physicians. However, with medical knowledge estimated to double approximately every 73 days^1^, educators, trainees, and students face mounting challenges in keeping pace with the expanding body of information. Medical students often struggle to synthesize the vast amount of material into cohesive, actionable knowledge^2^. Common study strategies include creating and reviewing flashcards and lecture handouts. However, student-made flashcards are often not updated annually^3^, leading to potential gaps in accuracy and relevance, while lecture handouts frequently include information that exceeds the scope required for medical students^4,5^. These limitations underscore the need for more efficient, targeted, and adaptable learning strategies.

Artificial intelligence (AI) has emerged as a transformative tool across several fields, including medicine. One rapidly advancing area within AI is the development of large language models (LLMs). These models are trained on extensive text corpora, enabling them to generate contextually relevant and sophisticated outputs. Initial applications of LLMs in medicine have demonstrated their utility in tasks such as automated clinical text summarization^6^, clinical decision support^7^, and writing patient clinic letters^8^. One particularly promising avenue for LLMs lies in medical education. With their capacity to process vast amounts of information and generate tailored content, LLMs are uniquely equipped to address the challenges of rapidly expanding medical knowledge. Medical students, in particular, stand to benefit significantly, as these models can streamline the synthesis of complex information, provide personalized study materials, and enhance understanding of intricate medical concepts^9–11^.

Despite the rapid evolution of AI technologies, research into their applications within medical education remains limited. Studies have evaluated the performance of LLMs on standardized exams^12–14^ and their ability to assess medical students during Objective Structured Clinical Examinations (OSCEs)^15^. However, few studies have explored how LLMs can be used to meaningfully and measurably enhance learning in medical education^16^.

In this study, we deployed an AI-based pipeline to generate lecture-specific summaries and Anki flashcards for medical students across two academic blocks. Our objective was to evaluate the effectiveness of these AI-generated materials in improving students’ understanding, retention, and overall performance. By analyzing their utility and reception, we aim to contribute to the growing body of evidence supporting the integration of AI tools in medical education.

## Methods

### Model Input and Calibration

The pipeline consisted of a two-step process: audio transcription, followed by LLM-based content synthesis and generation. Accurate transcription is necessary to achieve high-quality downstream results. OpenAI Whisper^17^ was selected for transcription based on prior evidence of strong performance in medical speech recognition and on benchmarking using human manually-generated transcripts of evaluation lectures (Table S1) ^18,19^. For downstream content generation, only the text of the transcription without generated metadata was used.

To develop a representative evaluation dataset, three lectures on diverse topics prepared by different faculty lecturers were used: Innate Immunity, Biochemical Energy States, and Teratogens. Because the pipeline accepts only audio context, we excluded lectures that rely heavily on visual content, such as pathology and radiology lectures.

Two leading LLMs at the time of experimentation were evaluated: GPT-4o and Claude 3.5-Sonnet. For each selected lecture, we created three prompts for summarization and three for flashcard generation. Each prompt-LLM combination was tested with and without access to faculty-defined learning objectives. Human-annotators assessed outputs on two main criteria: hallucinations and coverage of faculty-specified learning objectives. Flashcards were also assessed on appropriate information density (Figure 1).

**Figure 1:**
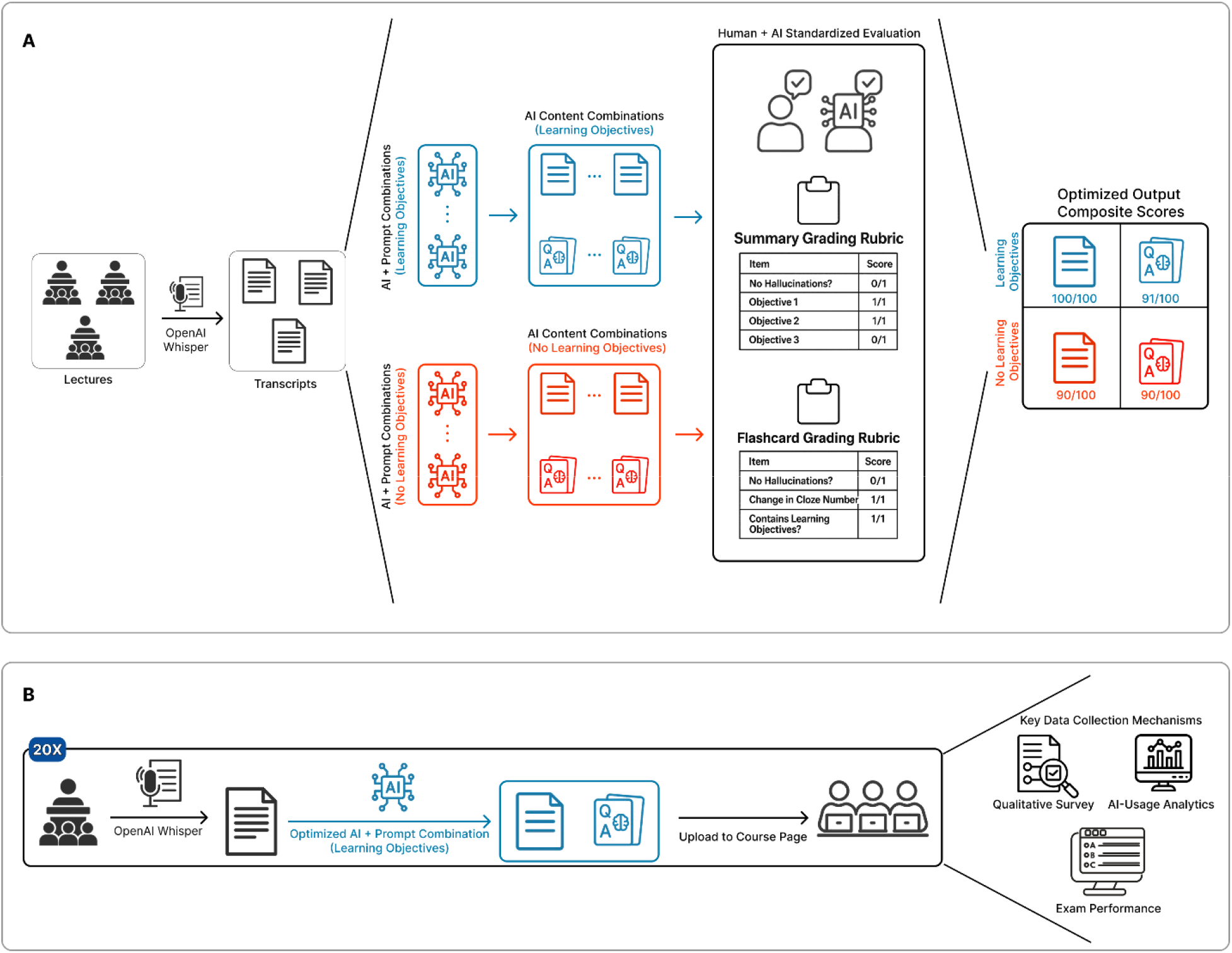
LLM Evaluation Framework, Content Generation, and Implementation Workflows. (A) Schematic depicting standardized human and AI calibration process to optimize AI model/prompt combinations for AI-generated flashcards and summaries. The composite score for AI-generated summaries is calculated as an equal weighting of the percentage of learning objectives covered and the percentage of outputs with hallucinations. For flashcards, the percentage of cards with a change in cloze-number (representing too much or too little information density per card) was also included in the calculation. (B) Process for application of optimized AI model/prompt combination to lectures (repeated 20 times, once per lecture) as well as associated student data collection.

We used GPT-4o to assess the adequacy of information density in each flashcard, given the inherent subjectivity in this task. The model determined whether each flashcard contained too little, sufficient, or too much information. This method of LLM self-assessment, central to agentic models, coupled with human evaluation, is supported in the literature^20–24^. This metric was used solely during prompt calibration, and not used to further edit the flashcards.

After collecting all manual and LLM-generated evaluations, a composite effectiveness score was calculated (ranging from 0 to 100) for each prompt-LLM combination, giving equal weight to all three evaluation categories: the percentage of learning objectives addressed, 100 -the percentage of hallucination-free outputs, and the percentage of flashcards with appropriate information density. The highest-scoring combinations for both flashcards and summaries were then used to generate the final study materials provided to students.

### Evaluation of LLM Usage on Student Performance

Institutional Review Board and institutional compliance approvals were obtained to ensure adherence to FERPA and other regulatory requirements. The study was conducted in the first-year pre-clerkship curriculum, in which students complete several blocks of interrelated basic science foundational subjects, such as Biochemistry, Genetics, and Pharmacology, within the Scientific Foundations of Medicine (SFM) course. The block spans 3-4 weeks of lectures from different courses (e.g., SFM, Pathology, Histology), and culminates in a comprehensive exam at the end of the block. For this study, we selected all 20 SFM lectures in two blocks, one focusing on genetics and the other on pharmacology. These subjects were chosen as initial candidates due to their minimal reliance on image-based knowledge, making them well-suited to a text-based learning approach ^25^. After each lecture, the pipeline described above was run (Figure 1). The resulting summaries and Anki flashcards were uploaded to the students’ course page on the learning management system, Canvas, which provides several metrics for file-level access, including file downloads and views. The lecture-hosting platform Panopto provided a commercial implementation of a default GPT summary (hereafter “Default GPT”), which served as a comparison. All AI-generated summaries and Anki flashcards were made available to the entire cohort of 143 first-year medical students through Canvas; engagement with the materials was opt-in, and usage was tracked at the file level via Canvas download and view analytics. To assess students’ subjective experiences, we administered a post-intervention survey at the end of Block 6, asking students to compare the AI-generated flashcards and summaries with existing study resources and to evaluate their perceived utility and time savings.

Exam questions were tagged by course and foundational subject. All exam questions were multiple-choice with a single best answer. Student performance was analyzed at the individual level across exam questions on pharmacology and genetics lectures. To account for baseline academic differences among students who self-selected to use AI resources and determine if AI resulted in the closing of any performance gaps, i.e., a “catch-up”, we incorporated prior academic performance from the three foundational preclerkship blocks preceding the intervention, being Cell Physiology, Immunology, and Biochemistry.

Performance data was linked to survey responses and usage metrics using de-identified IDs. The primary analytic cohort comprised 98 students with linked exam performance, Canvas usage, and post-intervention survey data, enabling within-subject integration of objective and subjective outcomes.

### Statistical Analysis

Paired t-tests were used to evaluate differences in transcript generation performance. Pearson’s r^2^ was used for correlation analyses related to increased usage and quantitative outcomes. Unpaired two-tailed Student’s t-tests were used to compare the performances of students who did and did not use AI materials. To test for equivalence rather than rely on the absence of a significant difference, we applied a two one-sided tests (TOST) procedure: equivalence between users and non-users was concluded only when the two-sided 95% confidence interval for the between-group difference fell entirely within a prespecified equivalence margin.

To determine whether subanalyses of AI content type and usage decisions were powered to detect meaningful differences, an alpha of 0.05, standard deviation of 10.78 (based on the previous year’s performance data for the same question categories), set equivalence margin, and the true allocation ratios between AI or no AI usage were used for two-sided calculations. The equivalence margin (representing the smallest educationally meaningful difference in exam performance) was prespecified as ±2 standard errors of measurement (SEM), corresponding to ±8.3 percentage points. SEM was estimated using a Cronbach’s alpha of 0.85, which is a standard value for medical school and standardized medical examinations ^26,27^.

For each student, a pre-intervention academic composite was calculated as the unweighted mean of the three prior block exam percent-correct scores. Baseline performance was then compared between AI users and non-users across each exposure group, including any AI use, summaries, and flashcards, within the pharmacology and genetics blocks. Comparisons were performed using Welch’s two-sample t-tests, with Cohen’s d reported as the standardized effect size.

Within-student performance change was calculated by subtracting each student’s prior three-exam mean from their intervention-block score. Differences in this change score between AI users and non-users were evaluated using Welch’s t-tests. Finally, we fit an ANCOVA model in which intervention-block percent-correct score was regressed on AI-use status, adjusting for the prior three-exam mean as a continuous covariate. Because change-score (difference-in-differences) and ANCOVA estimates rely on different assumptions about regression to the mean—the former assuming the two groups would otherwise change in parallel, the latter estimating the prior-to-outcome regression slope from the data—both were reported, and we inspected the within-group slope to quantify the degree of regression to the mean expected in the absence of any intervention effect.

All statistical analyses were performed with Python 3.10 using SciPy 1.15.1^28^. Qualitative feedback was also collected via free-text responses and thematically analyzed using GPT-4o.

## Results

### AI Output Validation

#### Transcript Accuracy

Two transcription systems (OpenAI Whisper and AWS Transcribe Medical) were evaluated on the training lectures (Table S1), using the original transcript, as well as a GPT-4o based self-corrected transcript. Word Error Rate (WER) was calculated relative to gold standard, annotator corrected transcripts. Word Error Rate with adjustment improvements for both OpenAI Whisper (WER=0.062, p=0.569) and AWS Transcribe Medical (WER=0.079, p=0.593) were neither statistically nor meaningfully significant. Across all lectures, the aggregation of OpenAI Whisper transcripts both with and without GPT-based corrections demonstrated significant improvement against an aggregation of AWS Transcribe Medical transcripts with an average Word Error Rate of 0.063 vs 0.081(paired t-test p=0.015). The GPT-corrected transcriptions did not demonstrate substantial improvement over the unmodified OpenAI Whisper transcripts.

#### Summary Prompt Validation

In total, 144 standardized criteria were manually assessed for 12 prompt-LLM combinations’ summary outputs across 3 lectures. The leading prompt-LLM combination demonstrated a hallucination rate of 0% and a mean representation of 100% of learning objectives for a total composite score of 100/100 (Table 1).

**Table 1.** Prompt Calibration Shows Substantial Learning Objective Coverage with Minimal.

| **Table 1. Prompt Calibration Shows Substantial Learning Objective Coverage with Minimal Hallucinations** |  |  |  |  |
| --- | --- | --- | --- | --- |
| **Prompt Type** | **Total Annotations Assessed** | **Learning Objective Coverage (%)** | **Hallucination Rate % (per output)** | **Composite Score** |
| **Flashcard Prompts** |  |  |  |  |
| **Objectives Provided** | **2038** | **100** | **4.8** | **91.3** |
| **Objectives Not Provided** | **2038** | **100** | **6.4** | **90.1** |
| **Summary Prompts** |  |  |  |  |
| **Objectives Provided** | **144** | **100** | **0** | **100** |
| **Objectives Not Provided** | **144** | **80** | **0** | **90** |

#### Flashcard Prompt Validation

In total, 4076 standardized criteria were assessed for 12 prompt-LLM combinations flashcard deck outputs across three lectures. The leading prompt-LLM combination demonstrated a hallucination rate of 1 hallucination per 21 flashcards (4.8%), a mean representation of 100% of learning objectives, and had 21.5% of flashcards recommended to be changed for suboptimal information density for a total composite score of 91.3/100 (Table 1). Within each lecture, the composite score was 86.7, 90.0, and 92.2 (each score was not weighted equally in the final calculation due to varying counts of learning objectives and flashcard outputs per lecture), demonstrating relatively consistent outputs and a narrow distribution across disparate inputs.

### Quantitative Outcomes Assessment

145 students in the pre-clerkship setting were considered initial candidates for the study, of whom 2 were excluded for opting out or failing to complete the curriculum. Exam performance data for 77 relevant questions was gathered from ExamSoft, and access and download analytics were collected via Canvas (**Figure 1B** and **Table 2**). Within the analytic sub-cohort of 98 students with linked exam, Canvas, and survey data, 92 students used the summaries for the Genetics block, and 26 students used the summaries for the pharmacology block. 60 students used the Anki Flashcards for Genetics, and 14 students used the Anki cards for the pharmacology block.

**Table 2.**
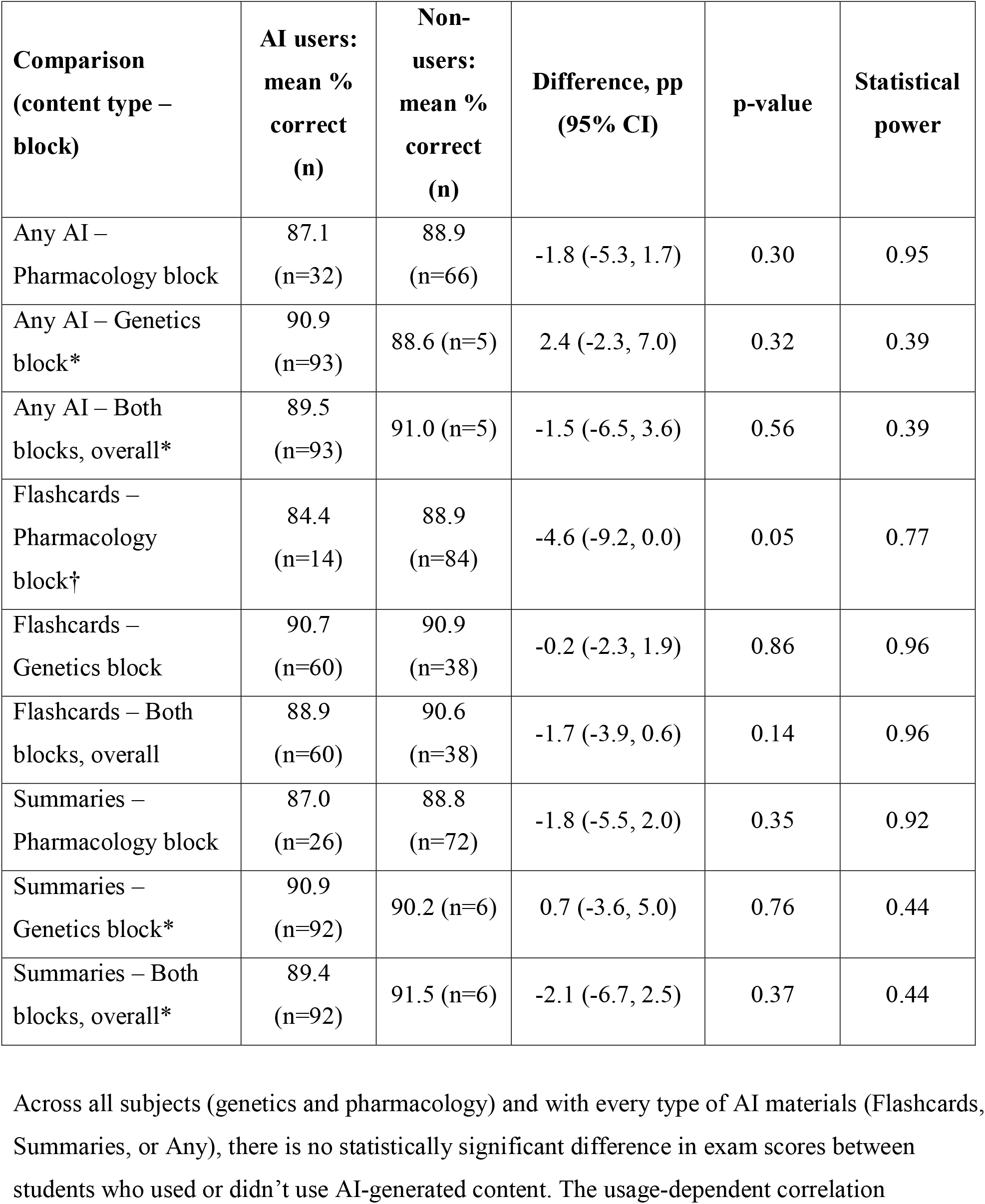
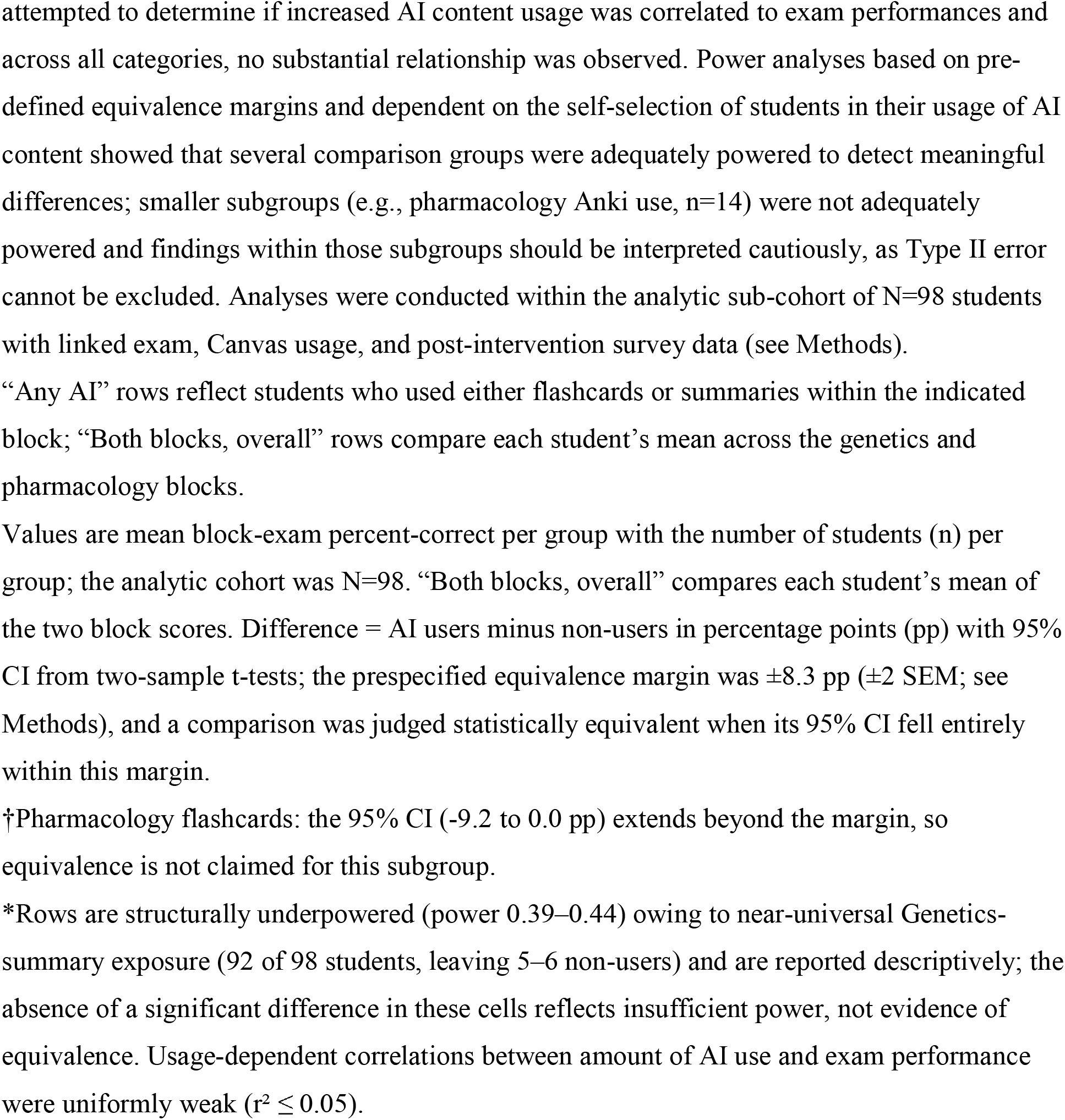
Block-Exam Performance for Users versus Non-Users of AI Content, by Content Type and Block.

Neither unpaired Student’s t-tests for exam scores for pharmacology or genetics for usage or non-usage of any type of AI material showed statistically significant differences (**Table 2** and **Figure 2**). To evaluate equivalence rather than the mere absence of a significant difference, the 95% confidence interval for each between-group difference was compared against the prespecified ±8.3 percentage-point margin (Table 2). Eight of the nine comparisons had confidence intervals lying entirely within this margin and therefore met the criterion for statistical equivalence. The single exception was pharmacology flashcards (difference -4.6 pp, 95% CI -9.2 to 0.0), whose interval extended beyond the margin, so equivalence was not concluded for this subgroup (n=14, power 0.77). The four comparisons defined by near-universal genetics-summary exposure (5-6 non-users; power 0.39-0.44) were structurally underpowered and are reported descriptively rather than as evidence of equivalence; non-significant findings in those subgroups may reflect Type II error rather than true non-difference (**Table 2**).

**Figure 2:**
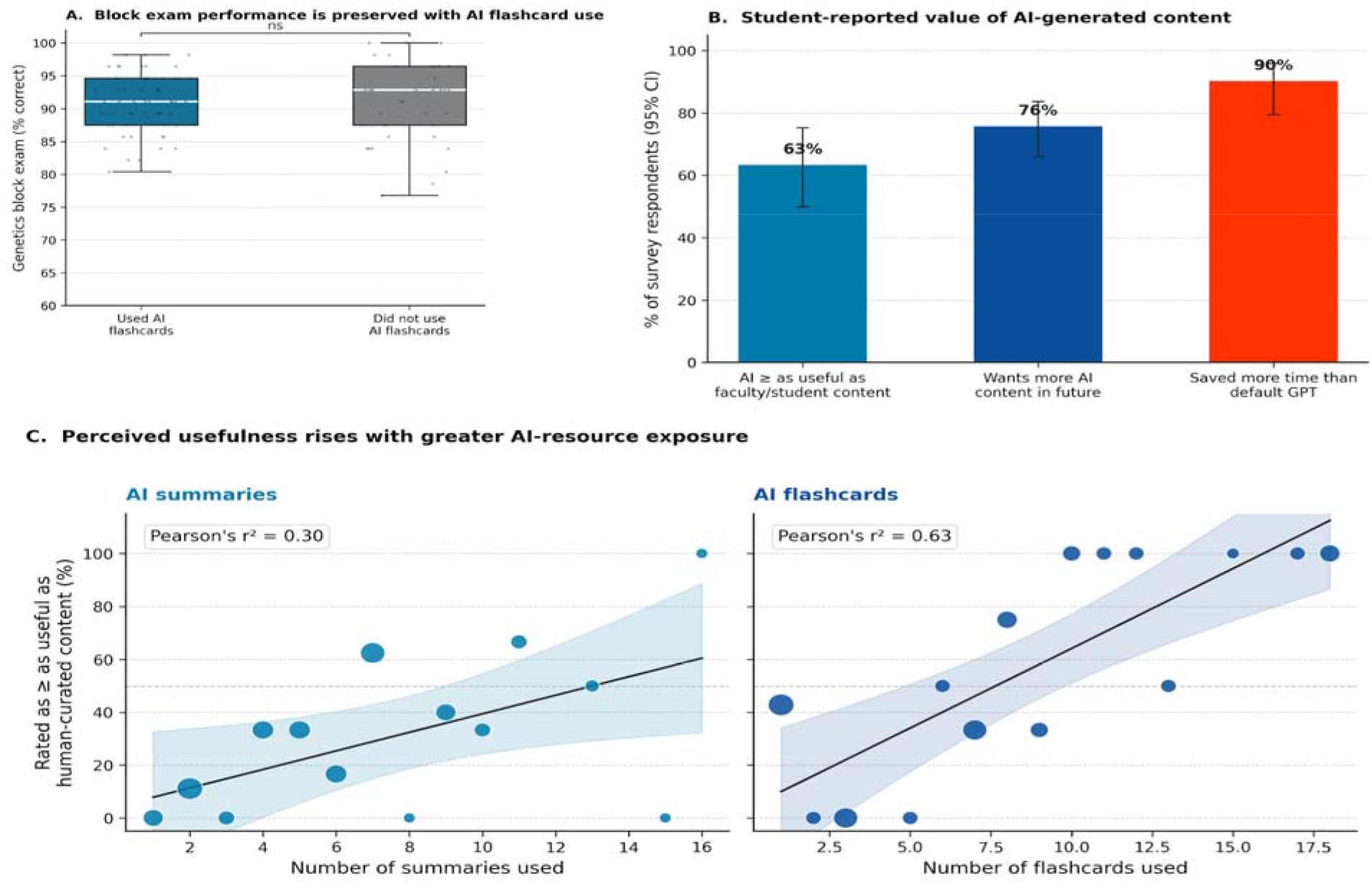
AI Study Content Usage Provides Key Perceived Benefits Without Compromising Exam Performance. (A) In genetics-associated questions, students who used AI-generated summaries had exam performance that did not differ significantly from students not using AI summaries. This trend was observed for all content types (genetics and pharmacology) and AI-generated materials (flashcards and summaries) (B) A majority of students indicated that AI-generated content was at least as useful as faculty or student-created content and would like to continue seeing AI-generated materials for future lectures. Over 90% of students using any customized study-generated AI content reported more time savings than the Default GPT summaries created by the lecture hosting platform. (C) As students used more AI-flashcard decks or Summaries, they were more likely to perceive it as equally or more useful to the analogous student or faculty-created content.

### Baseline Academic Comparison and Catch-up Analysis

Students who opted into AI-generated flashcards began with lower baseline (prior three-exam) performance than non-users, with medium effect sizes (pharmacology flashcards 83.4% vs. 88.9%, Cohen’s d = -0.59; genetics flashcards 87.1% vs. 89.7%, d = -0.36; pharmacology summaries 85.8% vs. 89.0%, d = -0.39), whereas the near-universally adopted genetics summaries (n=92 vs. 6 non-users) showed no baseline gap (d ≈ 0). Despite this disadvantage, AI users reached intervention-block scores that were no longer significantly different from non-users; for genetics flashcards, a 2.7-point baseline deficit narrowed to 0.2 points (difference-in-differences +2.5 points, p = 0.060). However, when analyzed by ANCOVA, the within-group prior-to-outcome slope was low (≈0.33), so after baseline adjustment for potential regression to the mean, the residual AI-use effect was small and non-significant (adjusted differences -2.6 to +2.2 points; all p > 0.25). Subgroups were also small (e.g., pharmacology flashcards, n = 14). These results are summarized in Figure 3 and Supplemental Figure S1.

**Figure 3:**
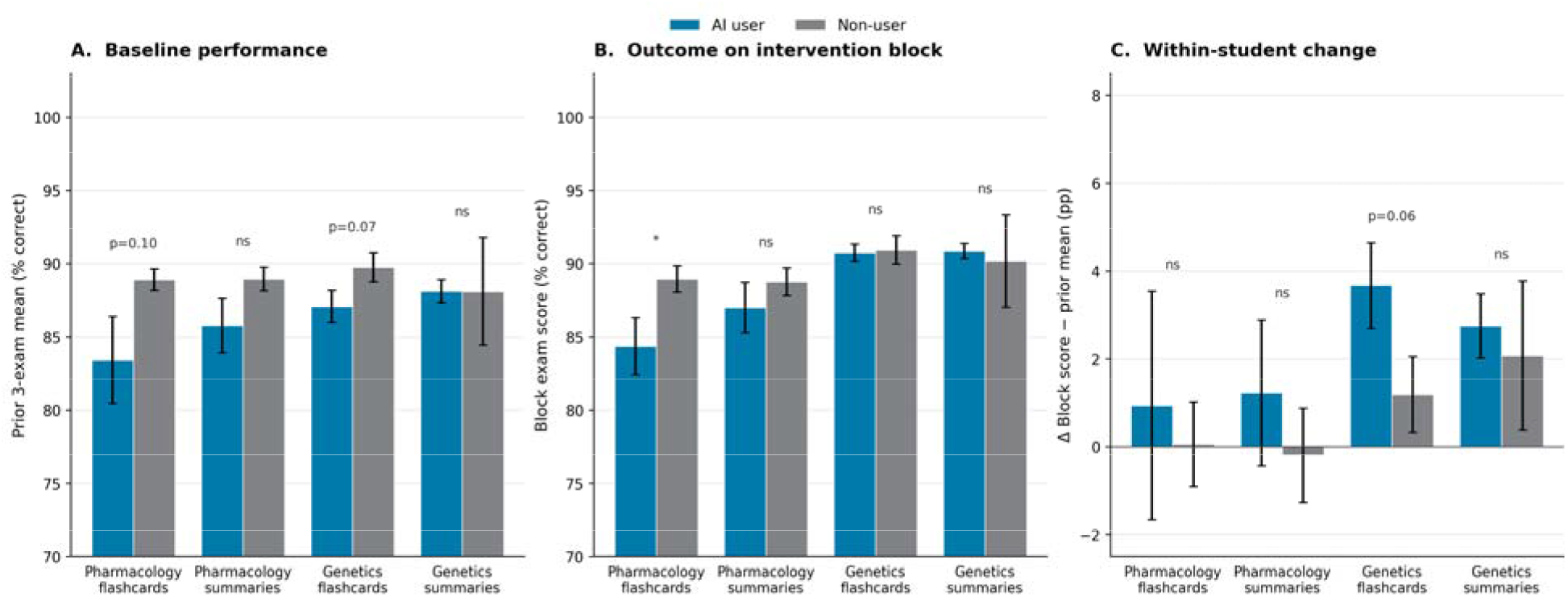
Baseline Academic Performance and Catch-up Pattern by AI-Resource Use. Comparison of pre-intervention academic performance (prior 3-exam composite: Biochemistry, Cell Physiology, Immunology) and intervention-block exam performance between students who chose to use AI-generated study materials and those who did not, stratified by content type (flashcards versus summaries) and intervention block (pharmacology versus genetics). (A) Baseline performance. Students who opted into AI-generated flashcards had numerically lower prior-exam composite scores than non-users (pharmacology flashcards p=0.10, genetics flashcards p=0.07). (B) Intervention-block outcome. AI users and non-users had block-exam scores that did not differ significantly despite the baseline gap. (C) Within-student change (block score minus prior 3-exam mean). The catch-up effect is most pronounced for the genetics flashcards (difference-in-differences +2.5 percentage points, p=0.06). Bars show group means; error bars are ±1 SEM. p-values are from Welch’s two-sample t-tests; an asterisk denotes p<0.05.

### Qualitative Outcomes Assessment

Of the 143 students enrolled, 98 completed the qualitative survey on AI-generated study materials. Results are summarized in Table 3. Notably, a majority of students reported that the AI-generated content was time-saving: 61% (95% CI [47%, 73%]) felt that the summaries saved them time while studying, and 74% (95% CI [59%, 85%]) reported the same for the Anki flashcards. In contrast, only 25% (95% CI [15%, 38%]) of students felt that the default GPT model saved them time, with less than 10% (95% CI [4.2%, 21%]) believing default GPT saved them more time than the AI-generated summaries (**Table 3 & Figure 2**).

**Table 3.**
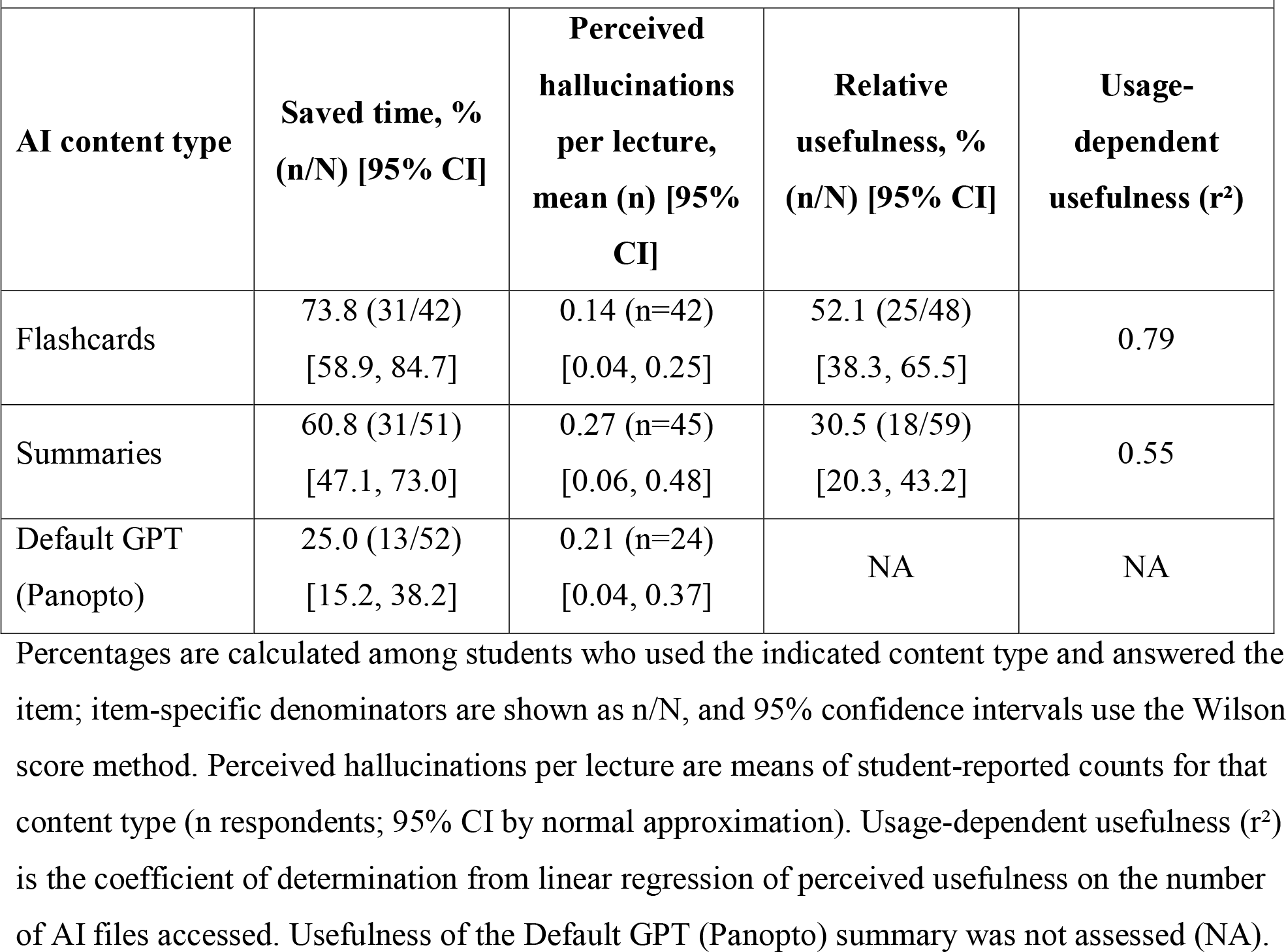
Student-Reported Time Savings, Perceived Hallucinations, and Usefulness of AI-Generated Materials.

| **AI-Generated Materials** |  |  |  |  |
| --- | --- | --- | --- | --- |
| **AI content type** | **Saved time, %**<br>**(n/N) [95% CI]** | **Perceived**<br>**hallucinations**<br>**per lecture,**<br>**mean (n) [95%**<br>**CI]** | **Relative**<br>**usefulness, %**<br>**(n/N) [95% CI]** | **Usage-**<br>**dependent**<br>**usefulness (r<sup>2</sup>)** |
| Flashcards | 73.8 (31/42)<br>[58.9, 84.7] | 0.14 (n=42)<br>[0.04, 0.25] | 52.1 (25/48)<br>[38.3, 65.5] | 0.79 |
| Summaries | 60.8 (31/51)<br>[47.1, 73.0] | 0.27 (n=45)<br>[0.06, 0.48] | 30.5 (18/59)<br>[20.3, 43.2] | 0.55 |
| Default GPT<br>(Panopto) | 25.0 (13/52)<br>[15.2, 38.2] | 0.21 (n=24)<br>[0.04, 0.37] | NA | NA |

Percentages are calculated among students who used the indicated content type and answered the item; item-specific denominators are shown as n/N, and 95% confidence intervals use the Wilson score method. Perceived hallucinations per lecture are means of student-reported counts for that content type (n respondents; 95% CI by normal approximation). Usage-dependent usefulness (r<sup>2</sup>) is the coefficient of determination from linear regression of perceived usefulness on the number of AI files accessed. Usefulness of the Default GPT (Panopto) summary was not assessed (NA).

Following the initial pilot across two pre-clerkship blocks, 76% of respondents expressed interest in continuing the AI-generated resource pilot in future blocks. Students reported using the summaries primarily for quick reference and orientation to lecture material, while the flashcards were more frequently used for active recall and general studying.

When comparing AI-generated materials to traditional resources, 64% (95% CI, [50%, 75%]) of students found that either AI-generated summaries or flashcards were as useful or more useful than the faculty-created summaries or student-created flashcards. Student perceptions of utility were associated with greater exposure: the more AI-generated files a student used, the more likely they were to rate the materials favorably. Moderate-to-strong positive correlations supported this relationship: Pearson’s r^2^ = 0.55 for summaries compared to faculty summaries, and a stronger r^2^ = 0.79 for flashcards compared to student decks (**Figure 2**).

Thematic analysis of the free-text responses demonstrated that over 50% of the comments were positive, often highlighting the convenience and clarity of the materials, while the remainder offered constructive suggestions. These included requests for more detailed content, visual aids, and improved integration with existing lecture materials.

## Discussion

This study presents an evaluation of optimized AI-generated learning content, specifically lecture summaries and Anki flashcards, deployed in a pre-clerkship medical education setting. Especially after the rise of virtual learning after the COVID-19 pandemic, the pre-clerkship setting continues to be study-content saturated while also placing a cognitive burden on students through the sheer number of learning resources within and beyond the formal curriculum^2,29,30^. This study sought to determine if AI could provide any benefits to trainee learning while also determining its utility in the broader setting of medical education.

We implemented a systematic comparison of multiple LLMs and prompt structures across diverse lecture topics and input configurations, with and without access to learning objectives. Each model’s outputs were evaluated for hallucination rates, coverage of learning objectives, and flashcard information density using a hybrid of human and LLM-based scoring methods. The use of straightforward, human-scored rubrics and composite scoring metrics ensured a high degree of rigor and reproducibility. Importantly, the final selected prompts demonstrated minimal hallucinations and near-complete coverage of faculty-specified content, key requirements for safe and effective deployment in educational contexts. Therefore, we believe we have produced a replicable blueprint for other institutions seeking to create high-quality, trustworthy AI-generated materials.

Qualitatively, students reported that the AI-generated materials offered meaningful benefits in terms of study efficiency and usability. A majority of students found the Anki flashcards (74%) and lecture summaries (61%) to be time-saving, markedly higher than the summaries provided by default GPT (10%). Students described using the summaries primarily for quick reference and orientation, while the flashcards supported active recall and broader content reinforcement. Thematic feedback also reflected appreciation for the clarity, conciseness, and organization of the materials, though students requested more detailed content, better integration with lectures, and the inclusion of visual aids.

Notably, students who used more AI-generated materials tended to rate them at least as helpful as pre-existing student or faculty-created materials (r² = 0.55 for summaries, r² = 0.79 for flashcards). While this suggests a relationship between use and perceived value, we cannot determine whether greater exposure increased satisfaction or whether students who already preferred the materials chose to use them more.

Quantitative results did not show a significant difference in exam performance across adequately powered comparisons. Students who used AI-generated summaries and flashcards achieved exam outcomes that did not differ significantly from their peers, falling within the prespecified equivalence margin in adequately powered comparisons, while reporting subjective time savings. AI-generated content did not raise exam performance beyond the existing curriculum, nor did it diminish performance in adequately powered comparisons. Well-calibrated LLM outputs may therefore represent an efficiency for programs producing study materials and an additional opt-in option for students, rather than a learning enhancement, in this resource-rich pre-clerkship environment. There is widespread concern that reliance on LLM-assisted medical learning will result in atrophy of skills and knowledge ^31^, but our findings indicate that when LLMs are appropriately implemented as a supplement to existing curricula, no measurable deficits were observed among learners who engage with LLM content.

Baseline comparisons using prior block-exam performance add context but should not be over-weighted. Across exposures, students who opted into AI-generated flashcards began with lower prior performance than non-users (Cohen’s d = -0.36 to -0.59), with the gap largest where uptake was lowest (pharmacology flashcards, 14 of 98; d = -0.59) and absent where uptake was near-universal (genetics summaries, 92 of 98; d ≈ 0). Despite this disadvantage, AI users reached intervention-block scores with no significant difference from non-users. A change-score analysis yields a marginal catch-up signal for genetics flashcards (+2.5 points, p = 0.06); however, this convergence could be attributable to regression to the mean, as after ANCOVA baseline adjustment the residual AI-use effect is small and non-significant (approximately +0.7 points, p = 0.46). Given self-selection, small subgroups, and a compressed scoring range, these data are suggestive of, but cannot establish, a catch-up effect and motivate future matched-cohort or randomized study.

There are limitations to our study. Unlike the clinical setting, the study resources available to pre-clerkship students are bountiful^2^, limiting our ability to examine the introduction of AI resources in a purely controlled manner. We were also limited by analytics on AI-material usage to downloads/views only, which did not provide a complete picture of the depth or consistency of resource engagement. Although several subgroups were adequately powered to detect meaningful score differences, self-selection into AI use led to uneven group sizes in some cases, which reduced power and complicates causal interpretation. The analytic cohort was further restricted to the 98 of 143 enrolled students with linked exam, Canvas, and survey data; this subset, selected by survey participation, may itself differ systematically from non-respondents, adding a second layer of self-selection.

Furthermore, prior-block exam performance remains an imperfect proxy for baseline academic ability and does not capture cumulative GPA, MCAT score, prior use of spaced-repetition tools, study habits, motivation, or other unmeasured factors that may influence both AI uptake and exam performance. Therefore, while the observed pattern is consistent with the possibility that some students used AI-generated materials to compensate for a baseline academic disadvantage, self-selection into AI use cannot be fully adjudicated by these analyses.

Power limitations were most pronounced in smaller or near-universal exposure subgroups. Smaller subgroups, notably the pharmacology Anki cohort (n = 14, power 0.77), were borderline-powered, and non-significant findings within those subsets should be interpreted with caution because Type II error cannot be excluded. In addition, four comparisons in Table 2 (Any-AI Genetics, Any-AI overall, Summaries Genetics, and Summaries overall) were structurally underpowered (power 0.39–0.44) because 92 of 98 students in the analytic cohort used the Genetics summaries, leaving only 5–6 non-users. For these cells, the absence of a statistically significant difference reflects insufficient power due to near-universal exposure rather than evidence of equivalence, and we report them descriptively.

This analysis was also conducted at a single institution and across a limited breadth of pre-clerkship topics. We therefore cannot assume that these findings generalize to other curricular environments, learner populations, or topics more reliant on multi-modal data, such as radiology and pathology. Matched-cohort or randomized designs in future work would more cleanly disentangle the effect of AI-generated study materials from the characteristics of students who choose to use them. Despite these confounders, many of which are inherent to real-world medical education studies, this study shows that students who chose to use AI-generated materials viewed these resources favorably and considered them comparable to expert human-produced content, with the added advantage of substantial time savings.

### Implications for Medical Education

In a rapidly evolving educational landscape, AI may help streamline content production for educators and offer students additional opt-in study options. The results of this study offer several important implications for the future of medical education, particularly in how LLMs can be integrated into pre-clerkship learning environments.

First, the positive student feedback suggests that AI-generated materials can enhance the learning experience by improving efficiency, lowering cognitive load, and offering flexible alternatives to traditional study methods while maintaining student exam performance. Students valued the clarity, organization, and accessibility of these tools, indicating a strong appetite for well-designed AI interventions.

Second, our results support the notion that calibrated LLM-generated educational content can serve as a viable supplement to human-created materials, particularly when built on validated prompts that minimize hallucinations and maximize alignment with learning objectives (the final prompts used to generate summaries and flashcards are provided in Supplementary Appendix S1). This creates opportunities for scalable content generation in settings where faculty time and instructional design resources are limited, such as newer or under-resourced medical schools, global health training programs, or continuing medical education.

Third, the strong correlation between increased exposure and perceived utility suggests that longitudinal integration of AI tools, rather than one-off deployments, may be critical for maximizing student benefit. Medical schools looking to implement AI-enhanced learning should consider not only the tools themselves but also how students are introduced to and trained to use them effectively over time.

Finally, while this study focused on pre-clerkship education, the framework developed here could be extended to other domains where information overload is common, such as clinical rotations, shelf exam preparation, residency training, and continuing medical education. Future studies should explore the role of AI in augmenting learning in these contexts and its potential to personalize content based on learner needs and knowledge gaps.

### Hallucinations

Learning objective, hallucination rates, and composite score (also includes appropriate information density for flashcards) for optimal prompts from prompt calibration process. Whether or not faculty-defined lecture learning objectives were pre-provided to the model, substantial coverage of learning objectives was achieved. Hallucination rates remained consistently low, with no hallucinations in summaries and 1 hallucination per 21 flashcards (4.8%) when objectives were provided and 1 hallucination per 16 flashcards (6.4%) when no objectives were available.

## Data Availability

The data analyzed for the present study are available from the corresponding author upon reasonable request.

---

**Supplemental Figure S1:**
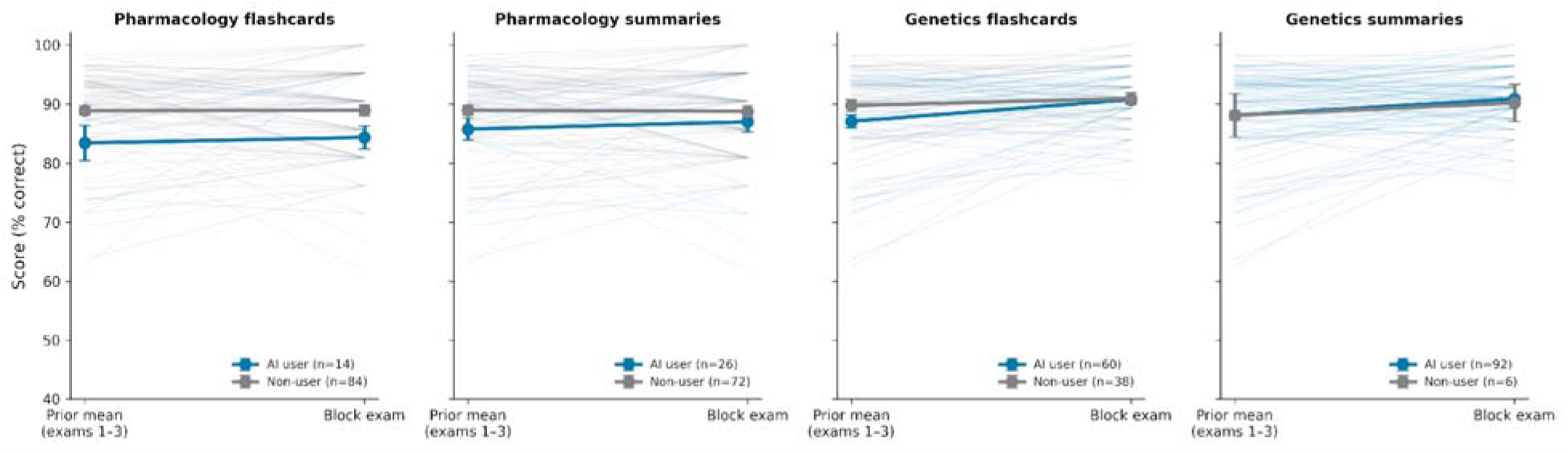
Per-Student Trajectories from Prior Exam Mean to Intervention-Block Exam. Individual-student slopes from the prior 3-exam composite to the intervention-block exam, stratified by AI-resource exposure and intervention block. Thin lines depict individual trajectories; bold markers depict group means ±1 SEM. The figure visualizes the catch-up pattern summarized in Figure 3, panel C.

**Table S1.**
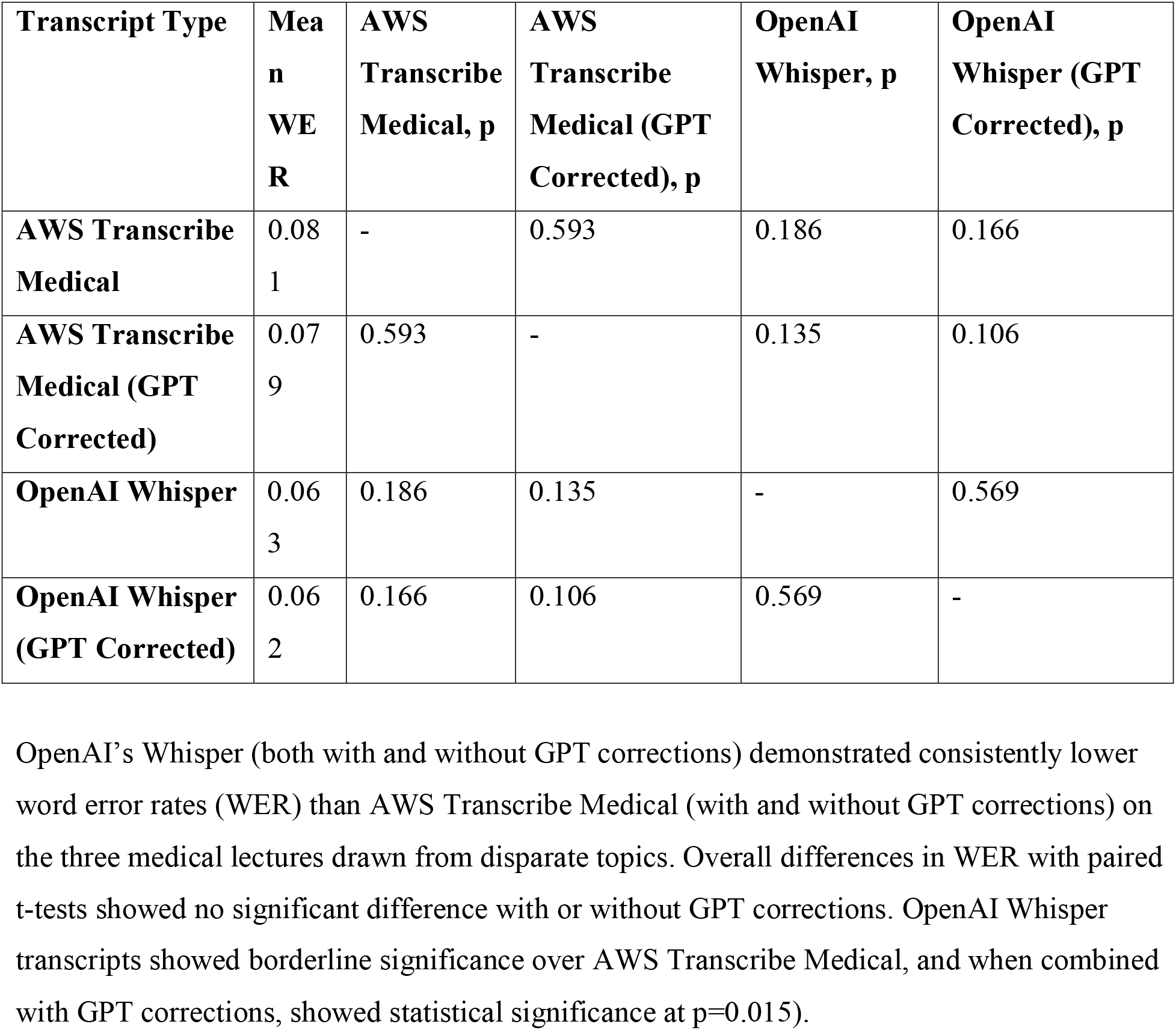
WER and Statistical Differences in Transcription Software on Medical Context.

## Supplementary Appendix S1

### Final Prompts Used for Summary and Flashcard Generation

The two prompts below are the final, content-agnostic prompts used unchanged across all 20 study lectures in the pre-clerkship blocks (Genetics and Pharmacology). The summary and flashcard prompts were each selected from the four prompt-configuration variants evaluated in the prompt-calibration experiments (Table 1, Methods) on the basis of highest composite score (combined hallucination, learning-objective coverage, and—for flashcards—information-density / cloze-number alignment). When faculty-supplied learning objectives were available for a given lecture, they were programmatically appended to the prompt before being sent to the LLM (see Methods, Section “Model Input and Calibration”).

#### S1.1 Summary Generation Prompt (verbatim)

> Summarize the lecture transcript provided, ensuring that all important points and details are captured in a concise and organized manner. Please make sure to include the learning objectives. This summary will be used by medical students for exam preparation, so include every key element needed for a thorough understanding without omitting essential information.

Runtime concatenation: when present, faculty-provided learning objectives were appended to the prompt on a new line. The transcript itself was passed as a separate message in the LLM API call. Model used in production: GPT-4o. Selection rationale: highest composite score in calibration (Table 1), including 100% coverage of faculty-defined learning objectives and 0% hallucination rate across evaluated lectures.

#### S1.2 Flashcard (Anki Cloze) Generation Prompt (verbatim)

> From the provided lecture notes and learning objectives, develop Anki cloze cards targeting the needs of a first-year medical student. Include key exam topics, important details, and foundational knowledge necessary for understanding the subject matter. Please generate however many cards is necessary for optimal retention. Please include an optimal number of cloze deletions.

Runtime concatenation: when present, faculty-provided learning objectives were appended to the prompt on a new line. The transcript itself was passed as a separate message in the LLM API call. Model used in production: GPT-4o. The output was post-processed into the .apkg format using the open-source genanki library before deployment to Canvas. Selection rationale: highest composite score in calibration (Table 1), with 4.8% per-card hallucination rate when objectives were provided and 100% learning-objective coverage.

## List of Abbreviations

AI: Artificial Intelligence
LLMs: Large Language Models
SFM: Scientific Foundations of Medicine (course)
WER: Word Error Rate
SEM: Standard Errors of Measurement
OSCEs: Objective Structured Clinical Examinations
IRBs: Institutional Review Boards
FERPA: Family Educational Rights and Privacy Act
GPT: Generative Pre-trained Transformer (used in model names like GPT-4o and Default GPT)
AWS: Amazon Web Services (from AWS Transcribe Medical)

## Declarations

### Ethics approval and consent to participate

The study protocol was reviewed and approved by the Brown University Institutional Review Board (IRB) (STUDY00000572) as a Quality Improvement initiative. The IRB determined that the project did not constitute Human Subjects Research and that informed consent was not required. The work was conducted in accordance with the Declaration of Helsinki and all applicable institutional policies, including those related to FERPA compliance, and was reviewed by the Office of the Registrar for compliance oversight.

### Consent for publication

Not applicable

### Availability of data and materials

Deidentified data used for analyses in this study are available upon request to the corresponding author.

### Author Contributions

J.K., H.Z., E.P., S.W., J.O., and T.S.M. contributed to the conception and methodology of the study, including its design. J.K., H.Z., J.O., and S.W. navigated compliance requirements and regulatory approvals. S.G., S.D.P., and S.E. were responsible for data labeling. J.K., H.Z., J.T., and H.M. conducted data collection and analysis. All authors participated in writing and editing the manuscript. S.W., T.S.M., H.M., and J.O. provided administrative support and facilitation throughout the study.

#### AI Usage Statement

OpenAI GPT models, Microsoft Copilot, and Grammarly were used in the initial editing of the manuscript with final revisions made by the co-authors. GPT Deep Research aided in collecting sources for the paper. GPT-4o was used to generate icons for Figure 1. GitHub Copilot assisted in drafting analysis code. All AI outputs were reviewed by authors for accuracy and originality. Authors affirm that the contents of this manuscript represents original work.

## Acknowledgements

The authors would like to thank Kyle Sloane and Anita Knopov MD, for assistance with data aggregation and collection, as well as staff from the Brown University Registrar’s office and Brown University and Brown University Health IRBs for feedback on study design and compliance.

